# Phylodynamics Of A Regional, Sars-Cov-2 Rapid Spreading Event In Colorado

**DOI:** 10.1101/2021.09.28.21262976

**Authors:** Kristen J. Wade, Samantha Tisa, Chloe Barrington, Kristy R. Crooks, Chris R. Gignoux, Austin T. Almand, J. Jordan Steel, John C. Sitko, Joseph W. Rohrer, Douglas P. Wickert, Erin A. Almand, David D. Pollock, Olivia S. Rissland

## Abstract

Since the initial reported discovery of SARS-CoV-2 in late 2019, genomic surveillance has been an important tool to understand its transmission and evolution. Here, we describe a case study of genomic sequencing of Colorado SARS-CoV-2 samples collected August through November 2020 at the University of Colorado Anschutz Medical campus in Aurora and the United States Air Force Academy in Colorado Springs. We obtained nearly complete sequences for 44 genomes, inferred ancestral sequences shared among these local samples, and used NextStrain variant and clade frequency monitoring in North America to place the Colorado sequences into their continental context. Furthermore, we describe genomic monitoring of a lineage that likely originated in the local Colorado Springs community and expanded rapidly over the course of two months in an outbreak within the well-controlled environment of the United States Air Force Academy. This variant contained a number of amino acid-altering mutations that may have contributed to its spread, but it appears to have been controlled using extensive contact tracing and strict quarantine protocols. The genome sequencing allowed validation of the transmission pathways inferred by the United States Air Force Academy and provides a window into the evolutionary process and transmission dynamics of a potentially dangerous but ultimately contained variant.

**SIGNIFICANCE:** SARS-CoV-2 spreads and mutates, negatively impacting containment. In this study, we use long-read sequencing to generate 44 SARS-CoV-2 genomes from COVID-19 patients associated with a rapid-spreading event on the USAFA campus, as well as a neighboring community for reference. We reconstruct the genomic and evolutionary signatures of the rapid-spreading event, and pin-point novel, protein-altering mutations that may have impacted viral fitness. These insights into viral evolutionary dynamics, in the context of contact tracing and a rigorous containment program, help to inform response efforts in the future.

## INTRODUCTION

The COVID-19 pandemic caused by SARS-CoV-2 has resulted in worldwide disruption and more than 4.4 million recorded deaths (Worldometers, August 22, 2021). Sequencing the SARS-CoV-2 genome from infected individuals is an effective means to track the dispersion and prevalence of the virus. Genomic tracking allows researchers to model viral evolution and identify possible variants of concern. For example, sequencing provided strong evidence that the virus was transmitted locally within Washington state as early as January 2020 (Holshue et al. 2020). Similarly, sequencing allowed public health officials to track the rise and spread of the highly infectious Delta variant, enabling more responsive policies (van Dorp et al. 2021). These sequencing efforts provide even greater power when coupled with viral evolutionary modelling (phylodynamics) in an epidemiological context (Rasmussen et al. 2011, 2014). This type of combined approach for tracking and predicting viral transmission is known as genomic surveillance and is a critical component of the modern public health response to viral epidemics (Pybus & Rambaut 2009; Mavian et al. 2020; van Dorp et al. 2021).

Due to the nature of the pandemic, many grass-roots sequencing efforts sprang up *de novo* around the world, leading to heterogeneity in sequencing quality and inconsistency in geographic sampling. For example, long-read Oxford Nanopore technology (ONT) with overlapping polymerase chain reaction (PCR) amplicons is commonly used to obtain sequences (Freed et al. 2020). Due to a relatively high error rate with ONT (in comparison with short-read sequencing technology), it can be difficult to accurately infer differences from the reference sequence (Oikonomopoulos et al. 2016; Buck et al. 2017). Because accumulated mutations are key data for inference of phylogenetics, convergence, and selected variants, it is important to be as confident as possible in mutational signatures. There has also been heterogeneity in sequencing between different areas. For instance, analyzing the GISAID database (Elbe & Buckland-Merrett 2017; Shu & McCauley 2017), there was little sequencing of Colorado genomes for much of 2020, making it challenging to understand the landscape of SARS-CoV-2 variant origins and evolution of local transmission during this phase of the pandemic. Finally, there also are conceptual challenges for this type of genomic surveillance due to incomplete knowledge of the etiology of epidemics, including stochastic environmental effects, sociological response, and phenotypic variance (Frost et al. 2015).

In the case study presented here, we focus on addressing some technical limitations in rapid SARS-CoV-2 genome sequencing to provide a snapshot of infection dynamics in Colorado in August to November 2020. We use replicate ONT sequencing and other experimental quality control steps to circumvent methodological pitfalls, describe the phylodynamics of a rapid viral spread event within a relatively controlled environment, and correlate it with complementary epidemiological data (Sitko et al., 2021). Sequence data was collected as RNA isolated from infected individuals sampled from two Colorado populations: samples collected for clinical testing at the University of Colorado (CU) Anschutz Medical Campus; and samples gathered predominantly from asymptomatic, randomly sampled individuals at the United States Air Force Academy (USAFA) who tested positive by PCR testing. Together, these samples represent a baseline for the two largest Colorado cities (Denver/Aurora and Colorado Springs) in late 2020, and document the rapid initial spread and subsequent containment of a highly-evolved variant.

## RESULTS

### Overview of SARS-CoV-2 sequencing and variant calling

We obtained 44 nearly-complete SARS-CoV-2 genomic sequences using RNA collected from anonymized individuals in two Colorado populations. The first was collected in August 2020 by the Colorado Center for Personalized Medicine Biobank at the University of Colorado (CU). The second was collected by the USAFA in September to November 2020. The CU samples were predominantly derived from clinical samples from Denver and Aurora obtained at the University of Colorado hospital, while the USAFA samples were predominantly asymptomatic, randomly sampled cadets with cases. In the case of the USAFA samples, because the testing and quarantine protocols established at the beginning of the school year resulted in a population free from COVID-19 when classes started in August (Sitko, et al. 2021), subsequent infections almost certainly originated from contact with the local Colorado Springs community.

We followed the ARTIC protocol to produce overlapping short ∼450 bp PCR-amplified segments (amplicons), using Minion sequencing and the Oxford Nanopore Technology (ONT) SARS-CoV-2 pipeline. Differences from the Wuhan reference genome sequence (NC_045512.2). were called as described in the methods to predict mutation events in the CU and USAFA genomes. We attempted to sequence 33 CU samples and 68 USAFA samples, of which ten and 41, respectively, did not amplify well enough to sequence, while five USAFA samples did not sequence well despite amplification. Ultimately, 44 SARS-CoV-2 genomes were fully and reliably sequenced with at least two replicates (Supplementary Table 1, 2).

We jointly analyzed the entire set of Colorado sequences and placed their plausible common ancestors on a phylogenetic network. In doing this, we identified two instances of convergent mutations. The mutation at position 14187 in genome J is shared with the descendants of ancestral node A12. However, J is a descendant of node A4, which is highly divergent from A12. Additionally, we identified that a mutation at position 24904 occurs in both genomes J and G. However, these two are separated by two ancestral nodes, and this mutation is not found in any other related genomes. Therefore, we conclude that these two mutations are chance convergent events.

We also discovered several putative mutation events that were incongruent among sequencing replicates based on sequencing coverage, genotype likelihood, or phylogenetic agreement. In one instance, genomes AR, AQ and AJ appeared to be missing calls for a mutation that should have been present, based on mutation content of closely related genomes of ancestor A3. However, this mutation, at position 27964, was called with very low-quality genotype scores in many other genomes. Therefore, we decided to infer that it should be present in these three genomes, but was simply missed in their replicates, as it occurred at a site that was difficult to identify.

In most other cases of mutation disagreement, we chose to be conservative and reject inconsistent mutations. For example, a pair of adjacent mutations at positions 24389-24390 were called in consensus sequences from 13 CU and USAFA samples but were only consistently replicated in two samples. The remaining 11 mutations were called in only one replicate per sample, and at generally lower-quality scores. Based on these incompatibilities, we excluded these mutation events from all genomes. Additionally, a series of polymorphisms from positions 19299 to 19550 were observed in several phylogenetically disparate genomes, but were not well replicated. These were also excluded. Another single nucleotide mutation at position 13094 appeared with low quality and inconsistent prevalence across replicates as well as at incongruent ancestral nodes and was therefore excluded. Finally, we excluded variants at positions 28882-28883 based on previous knowledge that these were problematic mutations (Kemp et al. 2021), although they would not have had a substantial impact on our phylogenetic network. These results underscore the importance of replicating sample sequencing and consideration of phylogenetic context whenever possible. The remaining mutation events were well replicated and placeable in a consistent phylogenetic network, and we concluded that they represent high-confidence sequences suitable for in-depth evolutionary analysis of SARS-CoV-2 infections in Colorado.

### Broad phylogenetic structure of the CU and USAFA genomes

All newly sequenced Colorado genomes appear to be descended from what we label the A1 ancestor, which is the likely ancestor of NextStrain’s clade 20A^1^. In other words, they all share four mutation events in common that separate them from the Wuhan reference sequence, at sites 3037, 14408, 241, and 23403. This result is not surprising, as this ancestor contains two amino acid-altering mutations in the RdRp and Spike proteins, which confer a competitive advantage over previous variants since its origin in January 2020 (Korber et al. 2020). Based on analysis using NextStrain (Hadfield et al. 2018), this variant’s early origin and competitive advantage over the original virus caused its descendants to represent 99% of genome sequences throughout the world as of August 2020, when the first of our samples were collected (Supplementary Figure 2).

14 of the 21 CU sequences and 75% of the USAFA sequences are descended from A2, which is itself a descendant of A1 and is the likely ancestor of NextStrain’s clade 20C, estimated to have originated in April 2020. A2 differs from A1 by two mutations, at sites 25563 and 1059, and rose to a highest frequency of 43% of sequenced genomes in North America by February, 2021. Thus, 20C for some time appeared well on its way to becoming the dominant variant in North America, and may have been at a competitive advantage compared to other early descendants of A1, but it has been losing ground to the well-documented Greek-letter variants (Alpha, Beta, Delta, etc.) since then (Figure 1) (Qiu et al. 2021; Washington et al. 2021; Weber et al. 2021). Because the Colorado A1 and A2 descendants (other than the Colorado Springs variant) are not highly clustered, the higher frequency of A2 in both CU and USAFA samples may indicate that the A2 variant was even more successful in Colorado than in the rest of North America.

**Figure 1.**
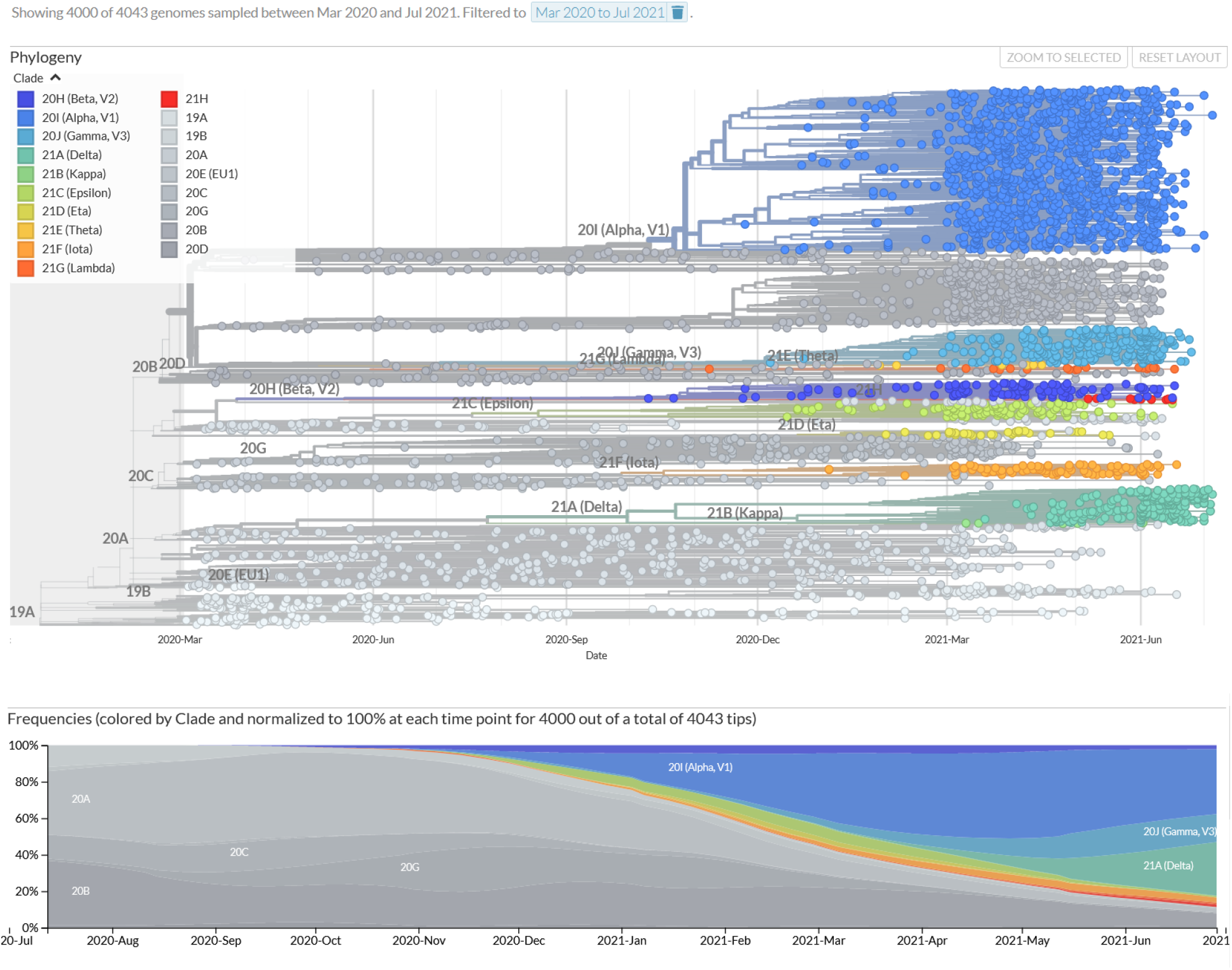
Eleven major SARS-CoV-2 clades in North America between March 2020 and June 2021, across 3923 genomes. Date accessed July 12, 2021.

We identified all plausible ancestral sequences in the Colorado phylogenetic network based on all observed different combinations of shared differences from the Wuhan reference sequence. This resulted in 13 inferred ancestors, which we label A1 to A13 (Figure 2, Figure 3, Supplementary Figure 3). We define all differences that can be parsimoniously mapped to branches of the network to be inferred lineage-defining mutations. There were 41 lineage-defining mutations and 135 novel mutations that were confidently identified from the network of 44 genomes that were sequenced (Table 1, Supplementary Table 3). Of the 41 lineage-defining mutations, 24 of these occurred in the ORF1AB gene, five in the S gene, five in the ORF3A gene, one in the ORF8 gene, and six in the N gene. 18 have documented amino acid substitutions already in the NextStrain database.

**Figure 2.**
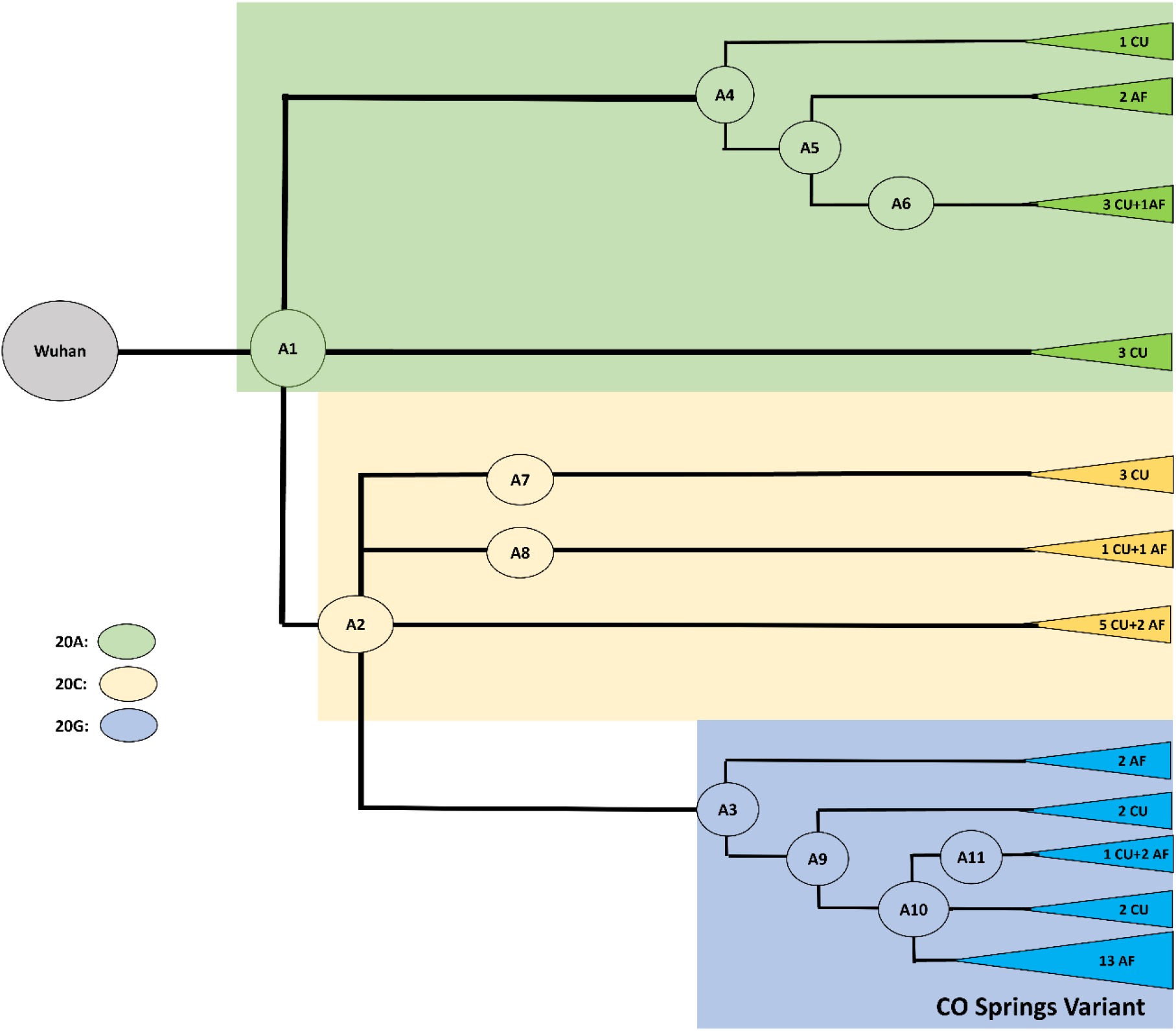
Evolutionary dynamics of the CUAF SARS-CoV-2 genomes. Tip labels indicate how many CU Anschutz (CU) and USAFA (AF) genomes are associated with ancestor. Color shading indicates which main phylogroup genomes belong to, A1-A3, corresponding to NextStrain clades 20A, 20C and 20G, respectively.

**Figure 3.**
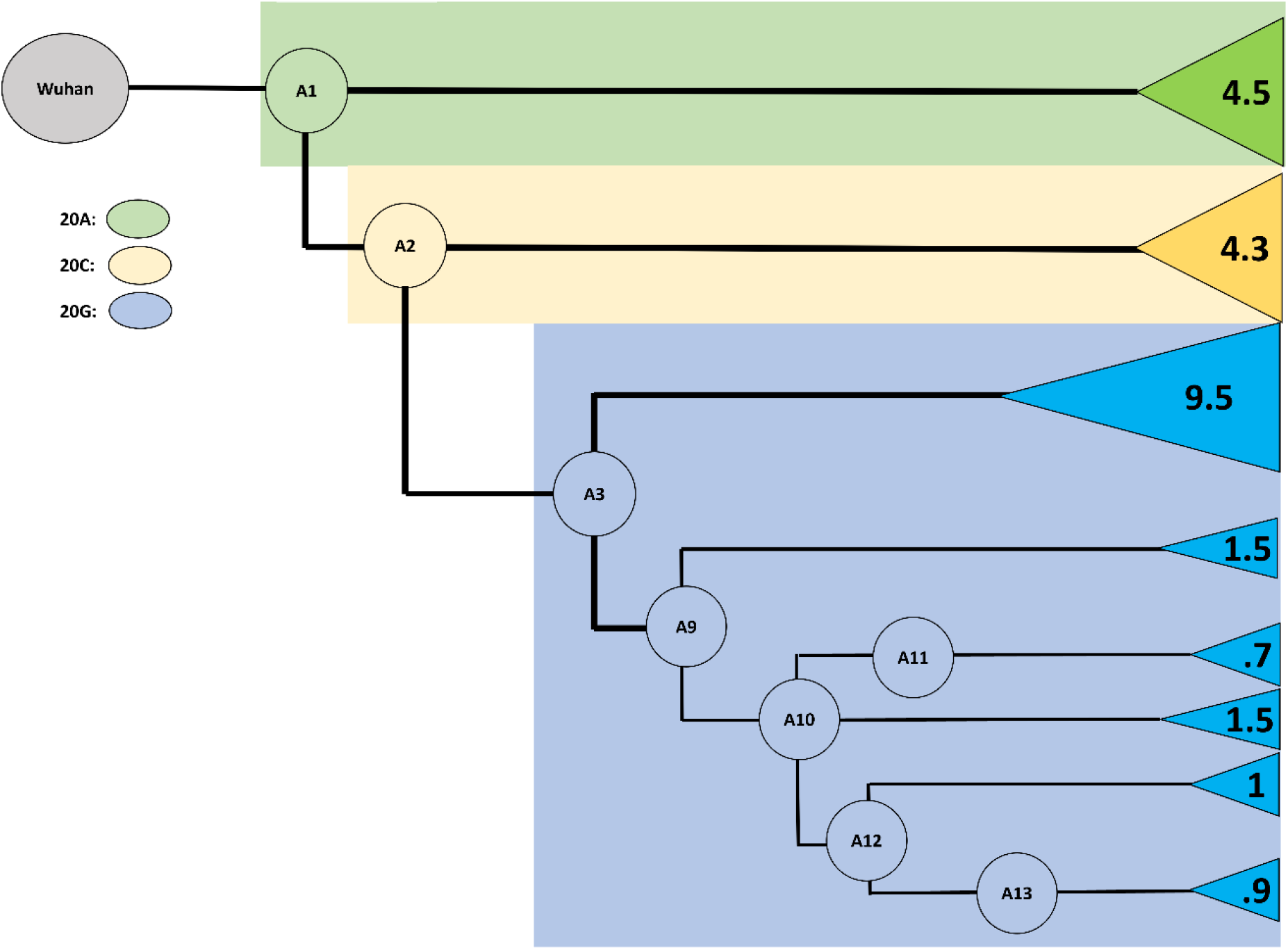
Closely related subset of Air Force samples suggests a novel, rapidly transmitting strain with low average mutation rate, the “Colorado Springs Variant” (A12-A13). Numbers at tips indicate average number of mutations per genome in each ancestral grouping.

**Table 1.**
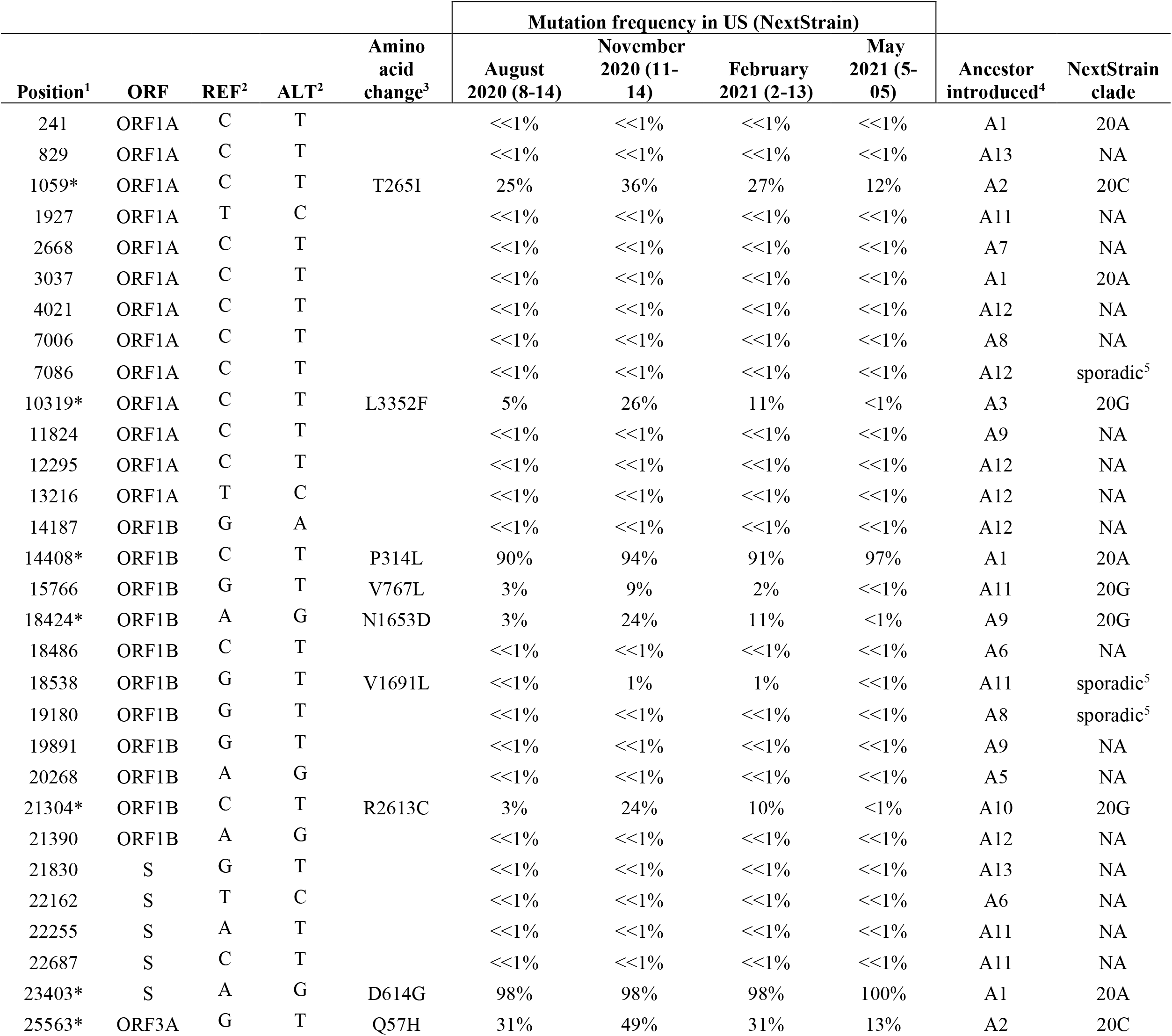
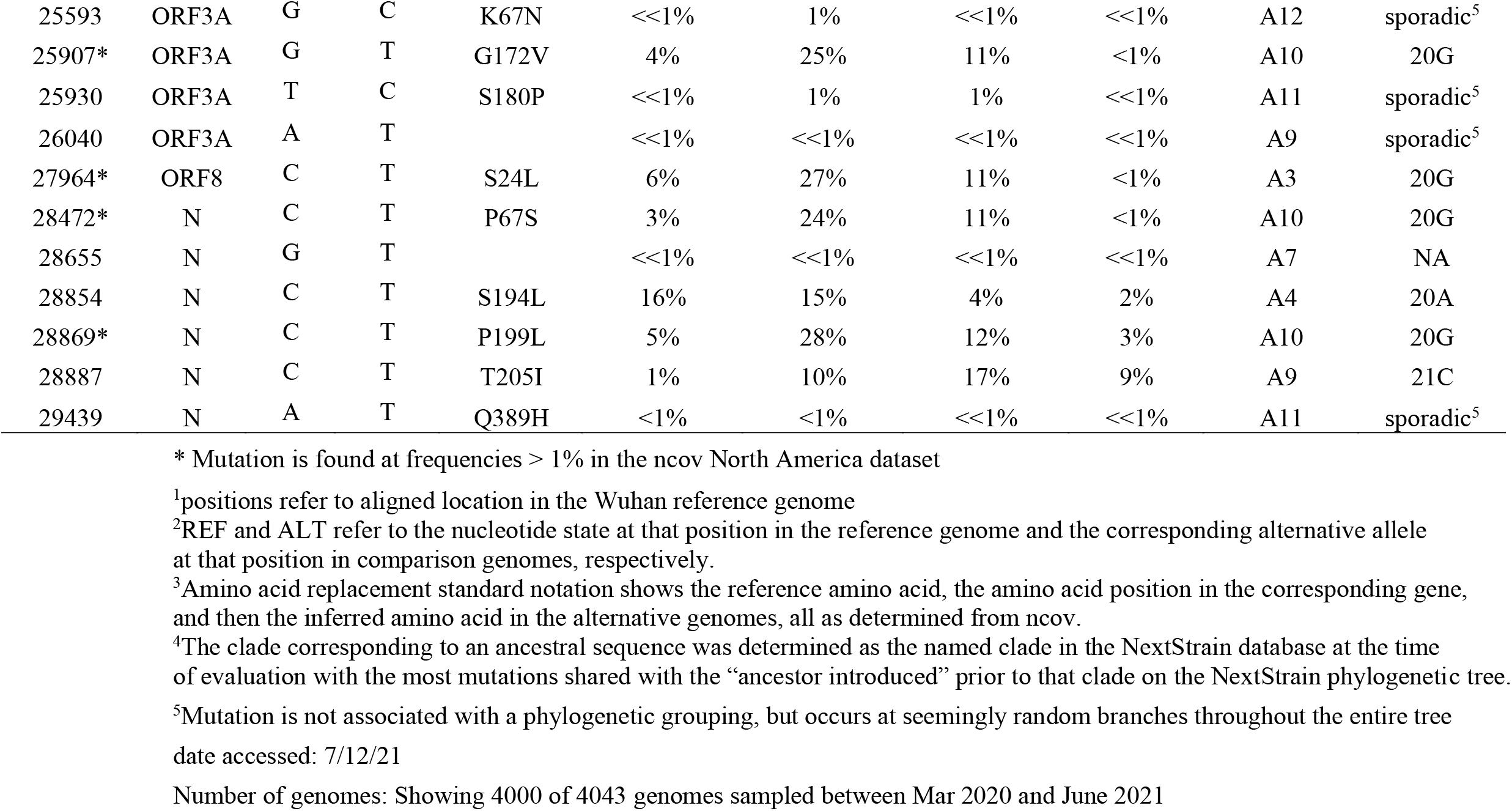
Lineage-defining mutations in a set of 44 Colorado SARS-CoV-2 genomes

These mutations were cross referenced against the NextStrain ncov database North American frequencies (henceforth, ncov) to evaluate their mutation frequencies and determine which of the eleven major clades, as defined by NextStrain at the time of evaluation, were represented in our sampling (Figure 2, Table 1). 18 mutations had no detectable frequency in ncov (≪1% in Table 1), and seven mutations were found sporadically across the ncov tree, with a prevalence of ∼1%. Of the 18 Colorado lineage-defining mutations that were more common in ncov, all were on the lineages leading to the series of ancestors A1, A2, or A3, corresponding to NextStrain clades 20A, 20C, and 20G. Based on shared mutations, 22 of the 44 Colorado genomes are descendants of A3 (and thus also A1 and A2), 12 are descendants of A2 (and thus also A1) but not descended from A3, while ten are descended from A1 but not from A2 or A3 (Figure 2). The inferred ancestors other than A1, A2 and A3 are organized such that there are three ancestors (A4-A6) descended from A1 but not A2 or A3, two ancestors (A7-A8) descended from A2 but not A3, and five ancestors (A9-A13) descended from A3. In this way, we were able to describe the patterns of relatedness and evolutionary dynamics between 44 Colorado SARS-CoV-2 genomes.

### Origins of a novel variant and rapid transmission event

Although many of the USAFA samples were likely picked up from the local Colorado Springs community based on detailed contact-tracing information and limits on off-base travel implemented by USAFA, a group of samples were also associated with a rapid-spreading event within the USAFA campus starting in late October 2020 (Sitko, et al., 2021). In looking at the distribution of inferred mutations on the Colorado phylogenetic network, we found that most common ancestors (A4, A5, A6, A8, A9) share a shallow network of divergence from the ancestors A1, A2, and A3, with one or two mutations separating each from an earlier ancestor (Figure 2, Supplementary Figure 3). Divergence from ancestors A1 and A2 involved an average of 4.5 and 4.25 mutations/genome (s.d. 2.5 and 3.5, range 1-9 and 0-11), respectively (Figure 3, Supplementary Table 4). Among the sampled sequences in this part of the tree, only two are identical. These results are in rough agreement with the idea that the mutations and most of the network diversification (other than A1, A2, and A3) were unselected and that mutations accumulated randomly with a rate for beta coronaviruses between 1.3 × 10^−4^ – 6.1 × 10^−4^ mutations per site per year (Drake & Holland 1999; Moya et al. 2004; Vijgen et al. 2006; Woo et al. 2012). A3, which corresponds to the common ancestor of NextStrain’s 20G clade, increased rapidly from August to November 2020 to a peak of 25% in North America (Table 1 Figure 4), and its dominant presence in the later USAFA samples may be due to stochastic events in the local community.

**Figure 4.**
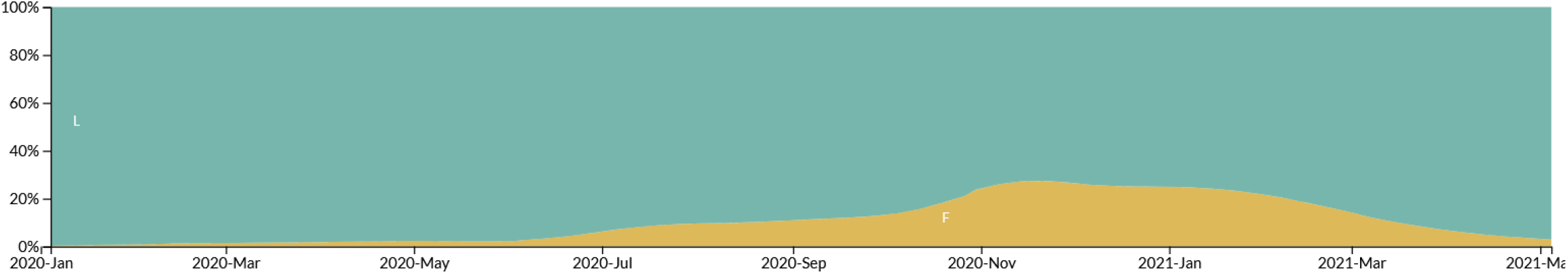
Frequency of a CUAF clade 20G mutation in North America between March 2020 and June 2021. The mutation pictured here is from ORF1A, genomic position 10319, amino acid position 3352.

The pattern in ancestors A10, A11, A12 and A13 is strikingly different. For example, divergence from ancestor A3 was much lower, with an average of only 1.77 mutations/genome (s.d. 2.9, range 0-12). This rate drops even further to <= 1 mutation/genome away from ancestors A11, A12 and A13 in the Colorado Springs variant lineage (Figure 3, Supplementary Table 4). Five mutations on the branch led to A10, and seven mutations on each of the branches led to two of its descendants, A11 and A12 (Figure 3). Furthermore, the highly derived A12 gave rise to 14 descendant sequences in the sample, including another shared ancestor differing by two mutations (A13). All of the descendant sequences from A12 and A13 differ from their ancestors by zero to three mutations, and there are respectively three and four sequences corresponding exactly to A13 and A14. The ancestors A10 and A11 have a few descendant sequences from CU individuals, indicating that they and A12 may have arisen from community spread. In contrast, the high prevalence of closely related USAFA samples in A12 and its descendants indicates sequence documentation of a rapid-spreading event, possibly involving one or more individuals. It is our inference that the source (ancestor A3) of the rapidly transmitting Colorado Springs variant (the descendants of ancestor A10) came from outside the campus originally.

Given the possibility that the Colorado Springs variant clade had some transmission advantage, we next considered the potential for the mutations to affect protein function. We evaluated the set of nine mutations that defined the CO Springs Variant ancestral clades A12–A13 (Table 2). These mutations have population frequencies <=1% in NextStrain (Table 1), and three result in non-synonymous amino acid substitutions. Two of these mutations (ORF1A/T2274I and ORF3A/K67N) arose on the branch leading to A12, the ancestor of the Colorado Springs variant. Based on literature review (see discussion), it is likely that all three mutations impact the biochemical properties of their associated protein and may have implications for viral fitness.

**Table 2.** Lineage-defining mutations contributing to the Colorado Springs variant (A12-A13).

| Ancestor | Mutation position | ORF | Protein | REF allele | ALT allele | Amino acid position | Codon change | Amino acid change | Biochem property change? |
| --- | --- | --- | --- | --- | --- | --- | --- | --- | --- |
| A13 | 829 | ORF1A | Nsp2 | C | T | 188 | AAC->AAT | N->N | N |
| A12 | 4021 | ORF1A | Macro Domain | C | T | 1252 | AAC->AAT | N->N | N |
| A12 | 7086 | ORF1A | Nsp3_C | C | T | 2274 | ACT->ATT | T->I | Y |
| A12 | 12295 | ORF1A | Nsp8 | C | T | 4010 | ACT->ACC | T->T | N |
| A12 | 13216 | ORF1A | Nsp10 | T | C | 4317 | GAT->GAC | D->D | N |
| A12 | 14187 | ORF1B | RdRp | G | A | 250 | AGG->AGA | R->R | N |
| A12 | 21390 | ORF1B | Methyltransferase | A | G | 244 | TTA->TTG | L->L | N |
| A13 | 21830 | S | Spike | G | T | 268 | GTT->TTT | V->F | Y |
| A12 | 25593 | ORF3a | Orf3a | G | C | 67 | AAG->AAC | K->N | Y |

### Genomic surveillance methods complement high-level contact tracing

From August 2020 through December 2020, USAFA utilized a random surveillance testing program, where a percentage of cadets (4-15%) were tested daily to identify asymptomatic or mildly symptomatic SARS-CoV-2 cases. All SARS-CoV-2 positive patients were interviewed, close-contacts identified, and class schedules reviewed to assess for additional contacts. These individuals were then placed into quarantine with testing and monitoring before release. If the individuals identified during contact tracing were not cadets, these close contacts were contacted following local public health guidance.

This method allowed for the identification of infection from a student, sports team, or community exposure. We were able to link multiple infections back to a rapid spreading event in late October 2021, which resulted in the strain clusters A12 and A13. Following the dramatic increase in infections, further lockdown measures were implemented at USAFA limiting new community introductions. At the end of the semester, a similarly rigorous testing and quarantine process occurred prior to release for the winter break, likely eliminating the strain from circulation within the population. The ability to contextualize genomic data with contact tracing information helps see a clearer picture for strain introduction, mutation, and propagation, while making assessment of subsequent viral fitness as SARS-CoV-2 continues to change.

## DISCUSSION

Our robust sequencing provides a snapshot of infections in Colorado in late summer and early fall 2020. There were many circulating variants in Colorado at this time, and their dynamics broadly reflect strain variation and divergence across the rest of the US. In addition, our analysis suggests that there were likely multiple introductions of SARS-CoV-2 into the USAFA cadet population, despite their restricted interaction with Colorado Springs. Most did not lead to subsequent outbreaks, and only one sustained a rapid evolutionary expansion, which we name the Colorado Springs variant. Prior sequences from the nearby Denver/Aurora area and pre-outbreak USAFA samples (likely reflecting the Colorado Spring community) indicate slow and potentially neutral evolution of variant twigs, which come from common ancestors of known expanding variants previously identified by NextStrain. The lineages immediately prior and adjacent to the Colorado Springs variant, in contrast, indicate bursts of evolution including amino acid altering mutations that may have affected its transmission properties. This variant may have been highly contagious, but its spread also appears to have been promoted by one or more rapid spreading events. Luckily, it appears to have been contained by the rigorous epidemiological control procedures (e.g. social distancing, mask wear, limited gathering, virtual learning) employed at the USAFA. The type of focused community-level data collected here may be key to understanding how SARS-CoV-2 spreads in local settings, which may often have highly idiosyncratic dynamics compared to the country as a whole.

We employed rigorous sequencing quality control and validation steps, including standard PCR and sequencing replicates of all samples, further replicates of any moderately ambiguous results, and comparison to evolving ancestral sequences as well as the standard Wuhan reference. This ultimately resulted in an inferred ancestral sequence network that contained only two convergent mutations and was parsimonious. Because we identified discrepancies that would have been accepted in less thorough (e.g. single-sequence replicate and single reference variant identification) protocols, we suspect, as do others, that an unknown number of variants in the GISAID database contain flawed sequences that may mislead phylogenetic and convergence analyses (Morel et al. 2021; McBroome et al. 2021; De Maio et al. 2021; Hodcroft et al. 2021).

The sequencing initiative presented here highlights the power of robust genomic surveillance to describe local viral dynamics, particularly when paired with epidemiological data collected with the patient samples. Sitko et al. collected these samples through a highly effective COVID-19 monitoring system at USAFA. Their approach relied on random sampling of individuals regardless of symptoms. In addition to their robust contact tracing steps, the USAFA was able to pinpoint their surge to a rapid spreading event on their campus in late October. This agrees with the conclusions described here, where we were able to phylogenetically reconstruct the divergence of these samples over the period of time from August to late November. Adopting this approach enabled the identification of the Colorado Springs variant in the context of enough branches to pinpoint a burst of change leading to the variant, and documented the spread of the variant to a large number of people over a short period of time. It is known that immunocompromised individuals can serve as accelerated cauldrons of intra-host viral evolution with selected and rapid accumulation of epistatically interacting mutations (Kemp et al. 2021), which might be an explanation for the burst of evolution we see here leading to the CO Springs variant. However, we do not know of such a case in the community, and the USAFA population is mostly young and extremely healthy, and the cadets likely interact with similarly young and healthy individuals in the local community. The possibility that rapid intra-host evolution could occur in such individuals, perhaps during long-term but largely asymptomatic infections, warrants consideration for further study.

It is important to track and model the evolution of highly adaptive strains that tend to rapidly rise in frequency in the population once they gain a sufficient foothold, but it is also important to describe patterns of viral evolution that may lead to attenuation. Such strains are likely to be found in sampling from asymptomatic patients because they tend to be less phenotypically severe cases. Attenuated strains have the potential to out-compete more severe strains due to the trade-off between virus transmissibility and severity (Armengaud et al. 2020). Studies such as the current one are well suited to capture a snapshot of this kind of variance, as samples were collected from both symptomatic and asymptomatic individuals, per USAFA’s randomized testing surveillance protocol (Sitko et al., 2021). Further, mutations in these strains can create reservoirs of mostly neutral mutations, possibly leading to gradual genetic drift over time (Fabre et al. 2012). In the event of a transmission bottleneck, variants could then rise to sustained, high frequency (Fabre et al. 2012; Munir & Cortey 2015). Such a scenario could contribute to antigenic shifts and viruses with a capacity to reduce vaccine efficacy (Armengaud et al. 2020). These scenarios, in which neutral variants propagate by chance, seem plausible as the default mode of spreading for SARS-CoV-2 (MacLean et al. 2021), punctuated by the rise of more transmissible variants of concern. While the Colorado Springs variant appears to have been confined to the isolated context in which it was found, similar variants may not be contained, and thus it is important to characterize them whenever possible.

The ancestor of the Colorado Springs variant (A12) contained two intriguing non-synonymous amino acid substitutions: ORF1A/T2274I and ORF3A/K67N. ORF1A/T2274I results in a shift from a polar uncharged to a non-polar residue at the third position of a three residue N-linked glycosylation site in the Nsp3 peptide. This type of post-translational modification to Nsp3 is thought to be important for insertion into the endoplasmic reticulum of host cells, though it is not known how a mutation at a glycosylation site would impact its ability to do so (Fung & Liu 2018). However, disruption of N-linked glycosylation on other viral peptides has been shown to be destabilizing and negatively impacts virus viability (Fung & Liu 2018; Dawood & Altobje 2020). The ORF3A/K67N mutation results in a change from a positively charged residue to an uncharged, polar residue. This particular residue occurs in an LKK peptide motif that was predicted to be a likely B-cell epitope by Azad and Khan, 2021. Because the mutation seems to increase the free energy of folding, it has the potential to alter a putative B-cell epitope, allowing the virus to better evade host immune responses (Azad & Khan 2021). Interestingly, the only recorded North American SARS-CoV-2 genomes containing this mutation are found in Mexico (Supplementary Figure 4). The NextStrain database (derived from the GISAID database) is a highly incomplete sample of existing viral strains, making it difficult to determine whether this mutation was imported to the Colorado area or arose *de novo* locally. The third amino acid altering mutation, along the lineage leading to A13, is located in the Spike protein at position 268. The change converts a small, hydrophobic valine residue to phenylalanine, which is also hydrophobic but contains a large six-carbon ring side chain. A shift in steric properties is likely to impact local structure, and thereby potentially modify protein function. Without further experimental studies, it is difficult to know how these mutations affect viral dynamics and the extent to which they enabled the rapid spreading event.

This case study, while limited in size and scope, is an exemplar to describe the viral phylodynamics of a locally confined rapidly-spreading transmission event, in combination with paired epidemiological data. Due to their rapid expansion, coupled with minimal mutation accumulation, rapid spread scenarios have little phylogenetic structure to describe (Leventhal et al. 2012), and the contact structures involved may strongly deviate from the average assumptions used in most epidemiological models (Frost et al. 2015; Pellis et al. 2015). We conclude that case studies similar to that presented here could assist in outbreak control, provide variant-origin replicates to obtain a broader view of the process and refine epidemiological models, and help in early detection and action against novel variants of concern when they occur in the future.

## METHODS

### Nanopore sequencing, alignment, and coverage assessment

Extracted RNA was obtained from either the University of Colorado BioBank (CU) or USAFA for samples collected between August and November 2020. Sequencing was performed according to the Nanopore Protocol for PCR tiling of SARS-CoV-2 (revision E, released Feb 6 2020) using the V3 primers (https://github.com/artic-network/artic-ncov2019/tree/master/primer_schemes/nCoV-2019/V3). 11 uL RNA was used for reverse transcription and initial amplicon PCR. Samples were processed in random order in two replicates. After the PCR and bead clean-up, samples were run on a 1.5% agarose gel, and those with visible bands at 400 bp were quantified using a Qubit fluorometer. Samples were end-prepped, barcoded and pooled together for sequencing. Samples were quantified using a Qubit fluorometer. The samples were then processed for downstream sequencing and analysis according to the Nanopore protocol. Sequencing was performed using R9.4.1 (FLO-MIN106D) Nanopore flow cell. Half of the prepared DNA library (7.5 ul) was diluted to a total volume of 12 ul prior to loading. A minimum of 40,000 reads was collected per barcoded sample. Samples with fewer than 40,000 reads were resequenced in a later run.

Sequenced reads for each barcode (corresponding to an individual sample) in fastq format were aligned to the reference Wuhan SARS-CoV-2 genome (NC_045512.2) using *mimimap2* (Li 2018). *Minimap2* parameters were run as follows for each barcode: -a –x splice –uf –k14 – secondary=no NC_045512.2.fasta. The resulting aligned reads were output into sam format and *samtools* (Li et al. 2009; Li 2011) was used to generate a binary alignment map (bam). Summary statistics describing read length, error rate, total number of reads, number of reads mapped and other quality metrics were generated using *samtools stats*. Further sequencing quality assessment was performed with *bedtools genomecov* (Quinlan & Hall 2010). For each barcode, a coverage histogram (-ibam), coverage map across consecutive intervals (-ibam –bga) and coverage map at each single nucleotide (-ibam –d) were generated. Finally, an R package (*minionCovidCoverage*.*R*) was used to visualize the genome-wide coverage distribution for each barcode individually. A custom python script (*findLowQualBases.py*) was used to obtain coordinates of low-quality genome sequence and stored in a bed file to mask the low-quality regions from the consensus sequence at a later step. Steps described here were called from a pipeline wrapper script, *runCovidSeqs.sh(Code available at: https://bitbucket.org/pollocklaboratory/covid19phylodynamicscode2021/)*.

Each individual genome included in further analysis was required to have a total read count greater than or equal to 40,000 reads, error rate less than or equal to 11%, greater than 95% of reads mapped to the reference genome, low quality sequence content less than 5% of the genome length, and low-quality runs at the 5’ and 3’ end of the genome not exceeding two hundred nucleotides in length. Any samples that failed to meet these thresholds were resequenced at least once.

### Variant calling, consensus sequence generation, and quality control

Variant likelihoods at each position were generated from the bam files of high-quality samples using the mpileup package of bcftools-1.11 (Li 2011), with the following parameters: -oU –d 200000. We then used bcftools call to make the variant calls, with respect to the reference genome (NC_045512.2), under the following setting: --ploidy 1 –vm –Oz. Variants assigned a ‘QUAL’ quality score <50 were masked out of the main analysis but stored separately should revision be required. Variants were stored in variant call format (vcf) and variant calls were mapped to NC_045512.2 with bcftools consensus. Following previous publications (Hourdel et al. 2020; Paden et al. 2020), positions with coverage less than or equal to 30X were marked as ‘N’ in the consensus genome sequence using bedtools maskfasta.

A negative control (water) and a positive control (SARS-CoV-2 RNA from ATCC) were included in each batch of samples. At least two sequencing replicates were performed for each sample to confirm that variants were reproducible; in some cases, especially for low concentration samples, samples were sequenced three or four times to improve call certainty. In later runs, to add further robustness against possible experimental artifacts, the two sequencing replicates were separated, and the order of samples was randomized within a batch. In the event of inconsistent variant calls among replicates, each single variant was scrutinized among all replicates and evaluated for genotype likelihood score (Li 2011), coverage depth at that position, and whether it occurred in other genomes. There were two instances in which a low-quality mutation (QUAL <50) was annotated: the mutation was called in all replicates or it was phylogenetically congruent with the other genomes present at a given ancestral node. Additionally, sometimes high-quality mutations did not fully replicate. To increase sensitivity, we annotated a mutation if it was called with a high quality in at least one replicate. To evaluate if primer error had contributed to recurring variant artifacts, primer sequences were mapped to all reads of a given barcode. The position of the primer match, relative to each read, was tracked and a distribution was created of all primer match positions across all reads (Supplementary Figure 1). The average matching read position of each primer was evaluated to assess whether any primers were enriched for mapping to ends of reads, or were distributed randomly as expected.

### SARS-CoV-2 phylogenetics

Plausible ancestors of the CU and USAFA genomes were inferred and compared to the NextStrain “ncov” database sample of 3983 genomes accessed on May 11, 2021 (Hadfield et al. 2018). Of these, 3923 were submitted in North America between March 5, 2020 and May 10, 2021, and will be referred to as North American NextStrain (NANS). For each mutation-derived sequence-altering event in both our newly inferred genomes and NANS, the estimated NANS frequency was evaluated at four different time points: August 14, 2020; November 14, 2020; February 13, 2021; and May 5, 2021. Each such event was also assigned to one of the eleven major NextStrain clades based on the ancestral context in which it first appeared.

Plausible common ancestors for all CU and USAFA sequences were independently inferred based on shared events deviating from NC_045512.2. These plausible ancestors were labelled A1-A13, with A1, A2 and A3 corresponding to NextStrain clusters 20A, 20C, and 20G, respectively. This understanding of the relatedness among genomes was used to assist in interpretation of uncertain variants, and some genome sequences with low-quality scores at a site in one or more replicates were assessed confidently if the variant in question was congruent with the ancestral predecessor. In so doing, we were able to confidently assign likely shared ancestors to all sequences, with only two apparent convergent events, and ignoring putative mutation events at 28881-28883, which are known to frequently recur as post in-situ (PCR or sequencing) errors (Kemp et al. 2021; de Maio et al., 2021 (website), de Maio et al., 2021 (website); McBroome et al. 2021; De Maio et al. 2021). Events separating ancestors were provisionally assigned based on differences between those ancestors to create an ancestral phylogenetic network, and non-ancestral mutation events were assigned to the branch leading to the individual sequences in which they occurred.

### Comparison to USAFA contact tracing and high-level contact tracing

A large number of the USAFA samples are descended from a series of closely related plausible common ancestors beginning with A12 that are mostly closely related to ancestor A3 (corresponding to NextStrain cluster 20G), but separated by nine mutation events mostly not seen elsewhere in the NextStrain database. This branch of the network appears to have been introduced into USAFA from contact with the local Colorado Springs community (Sitko, et al 2021), and we label it the Colorado Springs variant. The network relationships and identities among sequences were compared to USAFA contact tracing information (under USAFA IRB FAC20200035E) to examine consistency between our inferred genomic ancestral relationships and what was known about the contact path through the USAFA. The network was also used to examine whether the CO Springs variant arose from multiple community transfers or a single community transfer followed by spreading within the USAFA.

## Data Availability

All sequenced SARS-CoV-2 genomes are available on GISAID EpiCoV database, accession numbers available upon request. Relevant analysis code is on Pollock lab Bitbucket and publicly accessible.

https://bitbucket.org/pollocklaboratory/covid19phylodynamicscode2021/

## DATA AND CODE AVAILABILITY

All genome sequences were deposited in the GISAID database, accession numbers available upon request. All variant information and codes/script written specifically for this project are available at: https://bitbucket.org/pollocklaboratory/covid19phylodynamicscode2021/

## ETHICS STATEMENT

The United States Air Force Academy Institutional Review Board (IRB) determined the surveillance testing (FAC20200035E) was approved as Not Human Subjects Research in accordance with Title 32, Subtitle A, Chapter I, Subchapter M, Part 219: Protection of Human Subjects, Department of Defense Instruction 3216.02: Protection of Human Subjects and Adherence to Ethical Standards in Department of Defense-Conducted and –Supported Research and Air Force Instruction 40-402: Protection of Human Subjects in Biomedical and Behavioral Research.

Use of the University of Colorado Biobank samples for this study was reviewed by the COVID-19 Biobanking Committee on May 20, 2020 and approved May 22, 2020 in letter communicated by Matthew J. Steinbeiss, Special Projects Manager, Office of Regulatory Compliance, University of Colorado Denver, Anschutz Medical Campus, and signed by Thomas Flaig, Vice Chancellor for Research, University of Colorado Denver, Anschutz Medical Campus.

All samples were processed in the Rissland laboratory under IBC#1366. We were advised by Taylor Brumbelow (Human Research Protections, COMIRB Investigator Support, University of Colorado Denver, Anschutz Medical Campus) on May 19, 2020, that our research qualifies as “non-human subjects research” because we did not have access to or use identifiers such as exact dates of service or admission, county, or zip code.

## ACKNOWLEDGEMENTS

We thank members of the Rissland and Pollock labs for helpful discussions. KW is supported by NIH R01 GM083127. CB is supported by the T32 grant T32GM136444 awarded to the Molecular Biology graduate program. This work was supported by NIH grants R35GM128680 (OSR) and the RNA Bioscience Initiative.

## Footnotes

1 Although NextStrain calls its groupings “clades”, meaning they are collections of all viruses inferred to be descended from a single common ancestor, their tracking and labelling system confusingly goes against common usage of the word ‘clade’ by removing named clades from within larger clades, leading to a paraphyletic naming convention. Their naming conventions also do not track sequential origins of clades within clades, and they do not include many of the CO sequences. We prefer to explicitly track and label inferred ancestors and their relationships to CO descendants.

