## Supplementary Figures for "Phylodynamics Of A Regional, Sars-Cov-2 Rapid Spreading Event In Colorado"

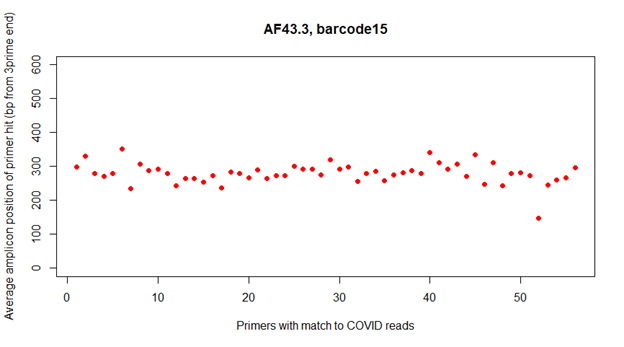

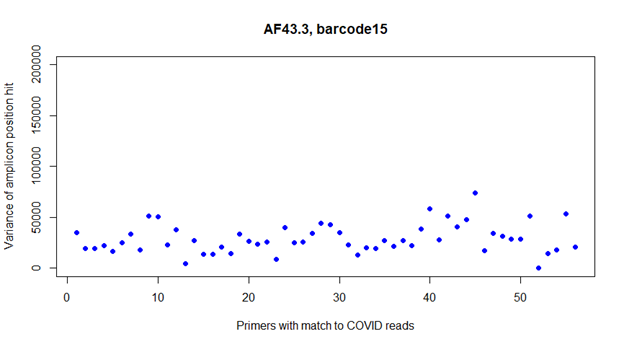

**Supplementary Figure 1**. Average matching amplicon position of sequencing primers. **1A** shows the average position along the yaxis, primer number along the x. **1B** shows the variance calculated for the distribution of matching positions for each primer.

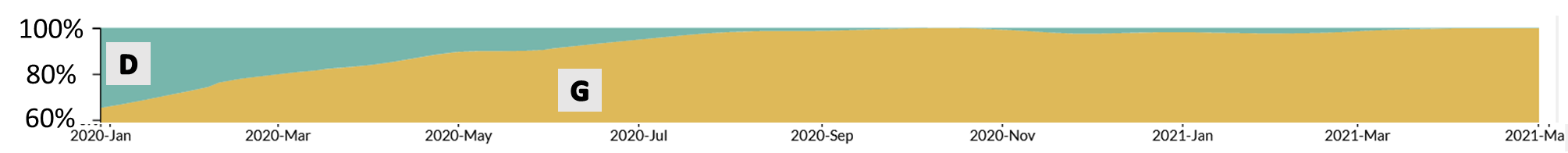

**Supplementary Figure 2**. Next Strain estimates of North American continental frequency of the D614G mutation, from March 2020-May 2021.

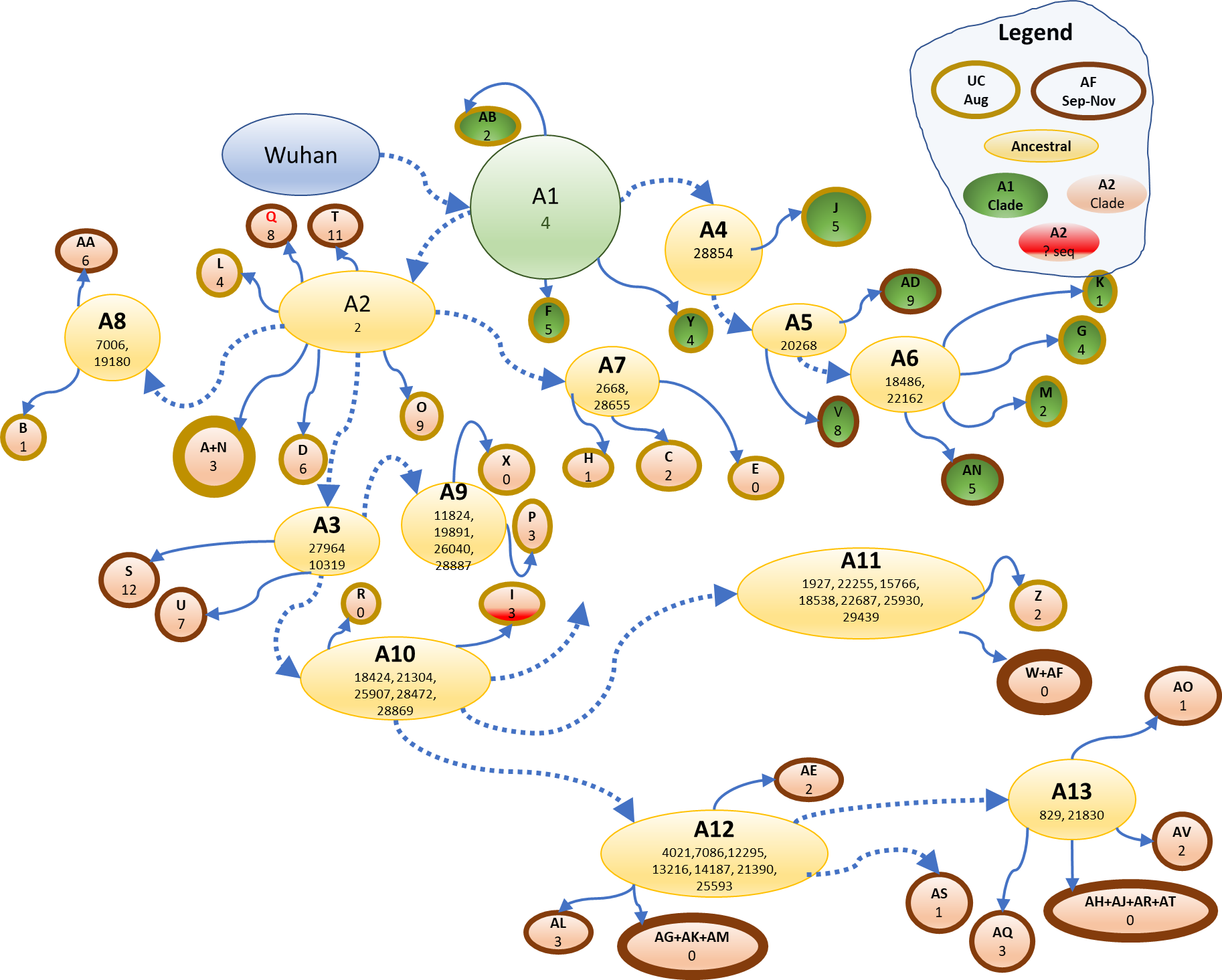

**Supplementary Figure 3.** Depiction of the ancestral network of 44 Colorado SARS-CoV-2 genomes. Each tip indicates the genome letter identifier and the number of mutations away from its most recent ancestral node. Ancestral nodes contain the name of the node (A1-A13) and the position of lineage-defining mutations which are inherited by all downstream lineages. Node coloring described in legend

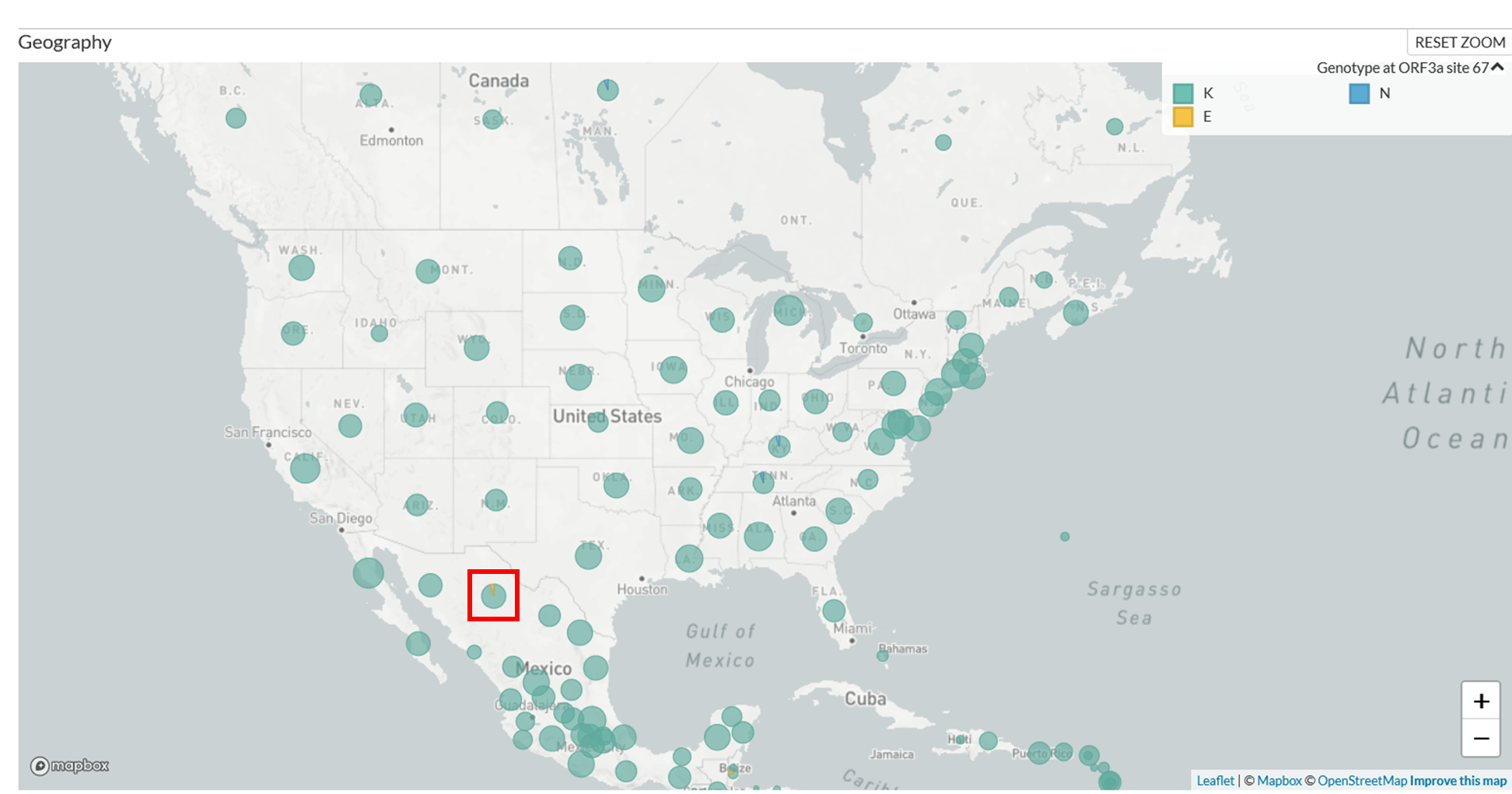

**Supplementary Figure 4.** Geographic location of the Orf3a K67N mutation in NextStrain. The only other documented instance of this variant in the NextStrain repository occurs in Northern Mexico.
